# Postoperative Cognitive Decline in Older Patients with Cardiovascular Disease and Preoperative Mild Cognitive Impairment

**DOI:** 10.64898/2026.06.16.26355755

**Authors:** María Mata, Magali González-Colaço Harmand, Pablo César Prada-Arrondo, Alberto Domínguez-Rodríguez, Jose Barroso, Iván Galtier

**Affiliations:** School of Psychology, Universidad de La Laguna (ULL), C/Profesor José Luis Moreno Becerra, s/n, Campus Guajara, 38200 San Cristóbal de La Laguna, Santa Cruz de Tenerife, Canary Islands, Spain; Head of Geriatric Medicine Department, Hospital Universitario de Canarias, Carretera Ofra, s/n, 38320 San Cristóbal de La Laguna, Santa Cruz de Tenerife, Canary Islands, Spain; Faculty of Health Sciences, Universidad Europea de Canarias (UEC), C/Inocencio García, 1, 38300 La Orotava, Santa Cruz de Tenerife, Canary Islands, Spain; Head of Cardiac Surgery Department, Hospital Universitario de Canarias, Carretera Ofra, s/n, 38320 San Cristóbal de La Laguna, Santa Cruz de Tenerife, Canary Islands, Spain; Department of Internal Medicine, Hospital Universitario de Canarias, Carretera Ofra, s/n, 38320 San Cristóbal de La Laguna, Santa Cruz de Tenerife, Canary Islands, Spain; Instituto de Investigación Sanitaria de Canarias, Carretera General La Cuesta, 75-77, 38320 San Cristóbal de La Laguna, Santa Cruz de Tenerife, Canary Islands, Spain; Centro de Investigación Biomédica en Enfermedades Cardiovasculares, Avenida Monforte de Lemos, 3-5, 28029, Madrid, Spain; Department of Psychology, Faculty of Health Sciences, Universidad Fernando Pessoa Canarias (UFPC), C/ La Juventud, s/n, 35450 Santa María de Guía, Las Palmas, Gran Canaria, Canary Islands, Spain; Instituto Universitario de Neurociencia (IUNE); Universidad de La Laguna (ULL), C/Profesor José Luis Moreno Becerra, s/n, Campus Guajara, 38200 San Cristóbal de La Laguna, Santa Cruz de Tenerife, Canary Islands, Spain

**Keywords:** Cardiovascular disease, Mild cognitive impairment, Cardiac surgery, Postoperative cognitive decline, Elderly

## Abstract

**Objective:** Older adults undergoing cardiac surgery may be vulnerable to postoperative cognitive decline. However, no studies have examined postoperative cognitive outcomes in older patients with cardiovascular disease (CVD) according to preoperative mild cognitive impairment (MCI). This study examined 12-month postoperative cognitive outcomes in older CVD patients according to preoperative MCI diagnosis and explored predictors of postoperative cognitive decline.

**Method:** Twenty-two older CVD patients (≥65 years) and twenty-five controls were included. Neuropsychological assessment was conducted at baseline in both groups and repeated 12 months after surgery in the CVD group. MCI was diagnosed using current clinical criteria. Postoperative cognitive change was examined across preoperative MCI groups.

**Results:** Fifty percent of patients met criteria for postoperative MCI, showing high diagnostic stability relative to preoperative frequency (45.5%). The preoperative CVD-MCI group showed a decline in working memory, executive functions, visual memory, and naming, whereas CVD-nMCI group declined only in verbal memory. Furthermore, CVD-MCI showed more heterogeneous postoperative cognitive trajectories of change than CVD-nMCI, who showed stability. Estimated IQ, APACHE-II score, and postoperative frailty were important variables in predicting the postoperative pattern.

**Conclusions:** MCI frequency remained high and stable in older CVD patients across the preoperative and one-year postoperative period. However, this apparent diagnostic stability masks subclinical cognitive decline, particularly among patients with preoperative MCI, who showed greater susceptibility to further impairment. Estimated IQ, APACHE-II score, and postoperative frailty may be considered relevant predictors of outcome. These results highlight the value of preoperative neuropsychological assessment for characterizing postoperative cognitive risk in older CVD patients.

## Introduction

Postoperative cognitive decline has been recognized as a significant complication in patients undergoing cardiac surgery [1,2]. In recent decades, the proportion of older patients (over 70 years old) undergoing cardiac surgery has increased [3,4]. At the same time, older age has been associated with a greater risk of neurological complications linked to surgery, including vascular damage (e.g. stroke), which can cause structural and functional alterations of the brain[5]. In addition, other complications such us postoperative delirium and postoperative cognitive dysfunction are common in older patients [6]. These findings suggest that older adults undergoing cardiac surgery may be particularly vulnerable to postoperative cognitive decline.

Vascular cognitive impairment refers to cognitive disorders, including mild cognitive impairment (MCI) and dementia, caused by cerebrovascular disease [7]. Although it has usually been related to acute vascular events (e.g. stroke and cerebral small-vessel disease), and has recently been associated with subclinical vascular diseases, such us cardiovascular diseases (CVD) [8].

CVD includes a group of disorders of the heart and blood vessels, often caused by accumulation of atheroma in the coronary arteries or heart valves, causing stenosis and reduced blood flow [9]. This chronic cardiovascular dysfunction can lead to a persistent reduction in systemic perfusion, which may cause cerebral hypoperfusion and, in the long term, diffuse cerebral ischemia. These disturbances in cerebral blood flow may contribute to brain alterations, and therefore to the development of vascular cognitive impairment. CVD has been recently associated with MCI before surgery, suggesting that postoperative cognitive decline may result in a context of preoperative cognitive vulnerability [10,11]. However, to our knowledge, no published studies have investigated longitudinal cognitive outcomes in patients with CVD in the presence of preoperative MCI.

Available studies have assessed pre- and postoperative cognitive impairment of CVD using screening tests or brief assessments with psychometric approaches without applying clinical criteria for MCI. Evidence from pre-post studies based on screening measures suggests an early decline in cognitive performance in the first days after surgery [12], followed by apparent recovery at medium-term of follow-up, a few months later [13,14], and a subsequent decline at longer-term follow-up, years after surgery [12,13]. Studies using brief cognitive assessments, have also described postoperative cognitive changes similar to those reported with screening assessments, in the early postoperative period and at follow-up intervals of less than one year [15–17]. However, a notable gap in the literature is the scarcity of studies that include long-term follow-up. To date, only one study has reported follow-up data of one year or more, finding postoperative cognitive decline in 42% of patients five years after surgery [18]. Nevertheless, none of the cited studies consider patients’ preoperative cognitive status using clinical criteria of MCI, a factor that may be important for understanding postoperative cognitive decline trajectories in CVD.

In addition to characterizing pre-postoperative cognitive changes, it is also important to identify the factors associated with postoperative cognitive decline in patients with CVD. The variables explored across studies are highly heterogeneous. Nevertheless, several sociodemographic (e.g. age, education level and intelligence quotient) and clinical factors (e.g. atrial fibrillation, biomarkers of heart failure, illness severity and frailty) have been proposed as relevant correlates of postoperative cognitive status [12,18–20]. However, to our knowledge, no published studies have examined predictors of different patterns of postoperative cognitive change.

Available studies about postoperative cognitive status in elderly patients with CVD have certain limitations: (a) to the best of the authors’ knowledge, no studies have reported preoperative cognitive status using clinical criteria for MCI while including postoperative follow-up assessments; (b) the timing of postoperative assessments is inconsistent across studies and is generally limited to early postoperative periods or to follow-up shorter than six months; (c) only three studies have reported follow-up data at one year or more after surgery, two of them based on screening assessments [12,13], and only one used a brief cognitive assessment [18]; (d) no published study has examined sociodemographic, perioperative and postoperative clinical variables as potential predictors of different patterns of postoperative cognitive change; and (e) notable heterogeneity can be observed across studies in terms of sample characteristics, with broad age ranges and the inclusion of middle-aged patients (around 50 years old).

Therefore, the aims of the present study were to: (a) examine changes in preoperative MCI status in older patients with CVD (≥65 years) and characterize their postoperative cognitive profile 12 months after cardiac surgery; and (b) explore whether sociodemographic, perioperative and postoperative clinical variables can predict postoperative cognitive decline.

## Materials and Methods

### Participants

The present longitudinal prospective study was conducted from 22 January 2018 to 22 January 2021. The baseline study included 34 patients with CVD and 25 healthy controls, all aged 65 years or older. CVD patients were diagnosed with coronary artery disease and/or heart valve disease and were scheduled to undergo cardiac surgery. Patients were recruited from University Hospital of the Canary Islands. The Healthy Control (HC) group included participants selected from the population-based sample of the Neuropsychological Studies Group of the Canary Islands [21]. Exclusion criteria for both groups were: a) prior diagnosis of dementia; b) major psychiatric disorder or intellectual disability; c) history of head injury with loss of consciousness; d) substance abuse; e) visual and/or hearing impairment that could interfere with neuropsychological assessment; and f) patients who were candidates for urgent or emergency cardiac surgery due to the inability to complete the assessment protocol. In order to ensure that HC participants did not have undiagnosed cardiovascular disease, their selection was based on medical history and absence of CVD symptoms. In addition, none of the HC participants met the diagnostic criteria for MCI [22]. Furthermore, they all had a score higher than 24 in Mini-Mental State Examination (MMSE) [23].

Twelve out of the 34 patients in the CVD group were lost to 12-month follow-up for various reasons: four due to COVID-19-related limitations, four declined further participation, two were excluded due to medical complications that interfered with the evaluation, one could not be contacted, and one had died. As a result, the final follow-up sample included 22 patients (Figure 1). Regarding the HC, the baseline measurements of the participants were used for the analyses, but this group does not have follow-up measurements.

**Figure 1.**
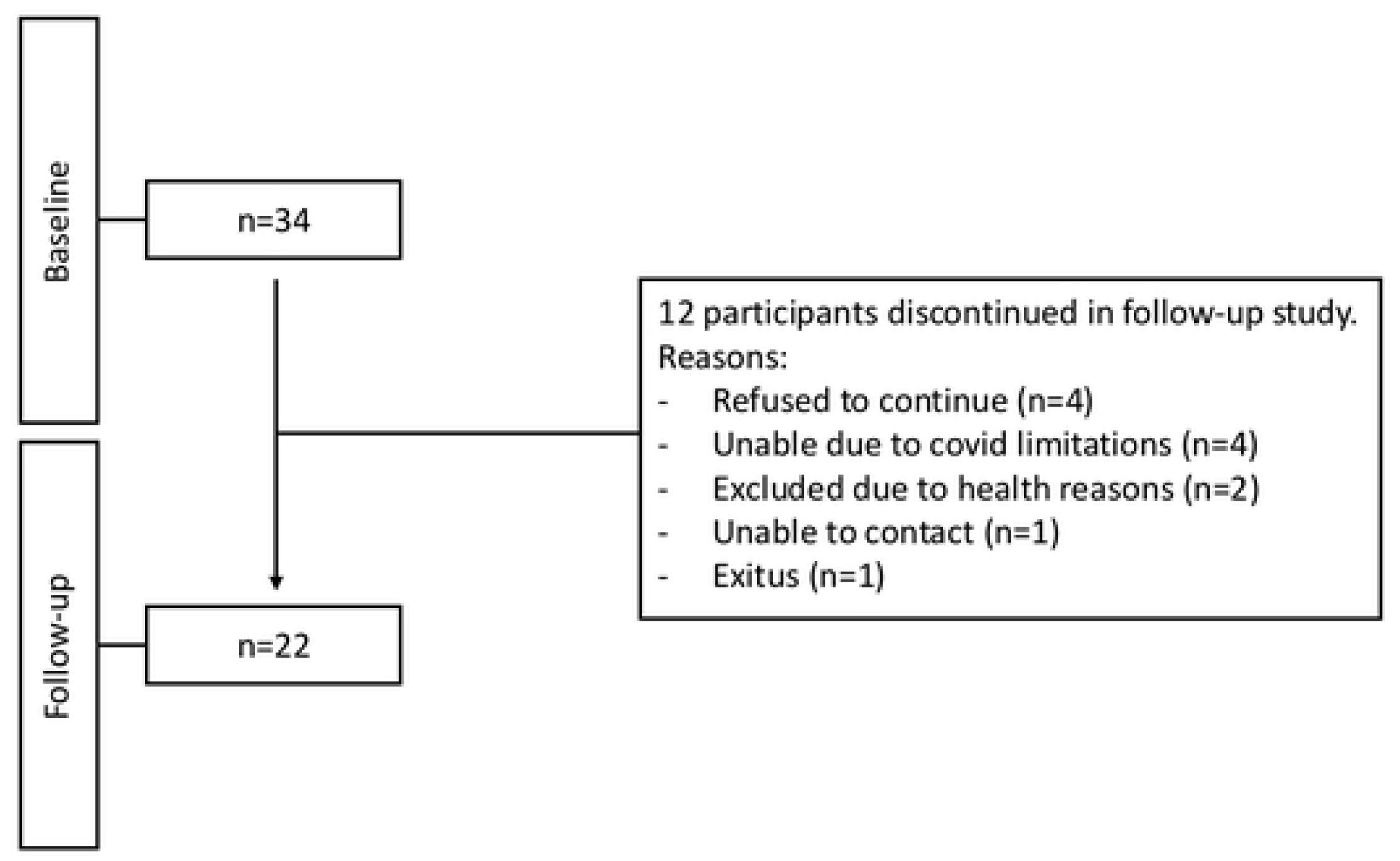
Reasons for discontinuing follow-up.

All participants were informed about the objectives of the study and gave written informed consent to participate voluntarily. The study protocol was approved by the Research Ethics Committee of the Hospital of the Canary Islands (reference: 2017_39 [CogCC], approved on May 25, 2017) and was conducted in accordance with the Declaration of Helsinki for research involving human subjects.

Both groups were matched at baseline in terms of age, years of education, sex, manual preference, and estimated intelligence quotient (IQ), assessed using the Information subtest of the Wechsler Adult Intelligence Scale–Third Edition, (WAIS–III) [24]. The Geriatrician Depression Scale (GDS) [25] was administered for the assessment of mood state (Table 1).

**Table 1.**
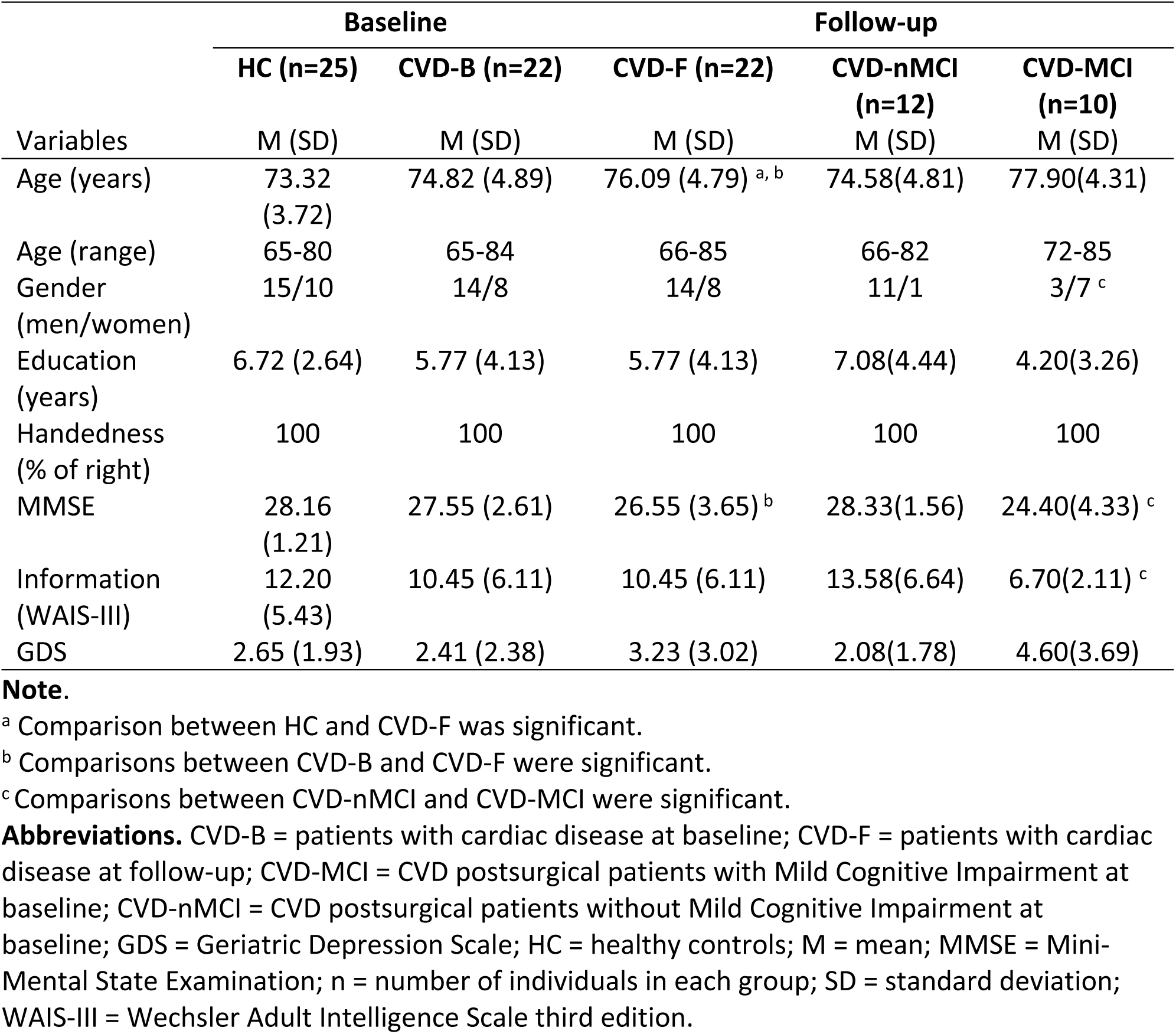
Sociodemographic and clinical characteristics of CVD patients and HC.

CVD patients were assessed at two time points: approximately 1 month before surgery and at 12 months after surgery. Both assessments were conducted by a multidisciplinary team consisting of a cardiologist, cardiac surgeon, geriatrician, and neuropsychologist. A thorough medical, physical, and neurological examination was carried out at each time point. The CVD patients and HC group underwent a comprehensive neuropsychological assessment that included two tests per cognitive domain: (1) attention and working memory were assessed using Paced Auditory Serial Addition Test (PASAT) [26] and Digit Span backward [27]; (2) executive functions were assessed using Verbal fluency tasks [28] and the Stroop Color–Word Test [29]; (3) learning and memory were assessed using the California Verbal Learning Test (CVLT) [30] and Visual reproduction [27]; (4) visuospatial functions were assessed using the Judgment of Line Orientation Test (JLOT, 15-item simplified version) [31] and Block Design subtest [24]; and (5) language was assessed with the Naming Test [32] and Center-embedded subordinate clauses test-comprehension test (CESCT), based on previous studies [33–35]. A detailed description of the test battery and administration procedures is available in the baseline research [10].

### Diagnosis of pre- and postoperative Mild Cognitive Impairment

The MCI diagnosis was established by a multidisciplinary team consisting of a geriatrician and neuropsychologists. The criteria defined by Winblad *et al.* were applied [22]. MCI diagnosis required evidence of cognitive decline, reported either by the patient and/or an informant, along with objective impairment on standardized cognitive measures. Impairment had to be evident in at least two tests, either within the same cognitive domain or across multiple domains. Cognitive impairment was defined as a score of 1.5 standard deviations or more below the mean performance of the HC group, in line with standard practice. In addition, patients had to preserve independence in basic activities of daily living, although some difficulties in instrumental activities were acceptable.

### Clinical variables as predictors of postoperative cognitive changes

A set of perioperative and postoperative variables was included in the analyses, according to the available literature to examine the clinical predictors of postoperative cognitive changes. The perioperative variables comprised: (1) postoperative atrial fibrillation, coded as a dichotomous variable based on its presence after surgery; (2) N-terminal pro-brain natriuretic peptide (NT-proBNP) levels, a biomarker of heart failure diagnosis and prognosis, dichotomized using a cutoff value of ≥ 5180 pg/ml as an indicator of severity; (3) and the Acute Physiology and Chronic Health Evaluation II (APACHE-II) score, a disease severity classification system for patients admitted to the intensive care unit, which was analyzed as both a continuous and dichotomous variable using a cutoff score of > 15 to indicate high mortality risk; and (4) postoperative frailty at 12 months was assessed using the Fried frailty phenotype [36]. Frailty was considered as both a continuous and dichotomous variable, with the dichotomous classification reflecting the absence (robust) or presence of frailty (prefrail or frail), based on a previous study [37].

### Statistical analysis

A nonparametric statistical approach was used because the Shapiro-Wilk W test showed that data deviated from a normal distribution.

Regarding the first aim of exploring the changes in cognitive status among the preoperative MCI subgroups (CVD-MCI and CVD-nMCI), three complementary analyses were performed. Firstly, the Wilcoxon signed-rank test was used to compare preoperative and postoperative performance in cognitive measures and the effect sizes were calculated. Secondly, a clinical approach was used to examine categorical changes in cognitive status across clinical subgroups. For each cognitive measure, the participant’s performance was classified at baseline and follow-up as either “normal” or “altered”. A score lower than 1 standard deviation below the baseline mean of the HC was considered altered. Based on this classification, each participant’s status on each cognitive measure was categorized as decline (change from normal at baseline to altered at follow-up), improvement (change from altered at baseline to normal at follow-up), or stable (no change in categorical status between assessments). The proportion of participants who changed cognitive status over time, for each cognitive measure, was analyzed. McNemar’s test was used to study the statistical significance of these categorical changes within clinical subgroups. Thirdly, an intrasubject approach was used to analyze overall patterns of cognitive change across measures. Each participant was classified in a “decline”, “improvement”, “mixed” or “stable” pattern, based on the number and direction of change in cognitive variables between baseline and follow-up. A “decline” pattern was assigned when the participant showed decline in three or more cognitive variables, and the number of declines was higher than the number of improvements. An “improvement” pattern was assigned when the participant showed improvement in three or more variables, and the number of improvements was higher than the number of declines. A “mixed” pattern was used when the participant showed at least two variables with decline and at least two with improvement, and the total number of declines and improvements was the same or differed by only one. A “stable” pattern was defined as having 10 or more variables without change and no more than two variables showing either decline or improvement (Table 2).

**Table 2.** Criteria for classifying patterns of cognitive change.

| **Cognitive change patterns** | **Numbers of variables with changes** | **Predominance in the direction of change** |
| --- | --- | --- |
| Decline | ≥ 3 with decline | No. with declines > No. with improvements |
| Improvement | ≥ 3 with improvement | No. with declines < No. with improvements |
| Mixed | ≥ 2 with decline and ≥ 2 with improvements | No. with declines = No. with improvements* |
| Stable | ≥ 10 without changes and ≤ 2 variables with decline or improvements | No. with stables > No. with declines or improvements |

**Note.**
\*Or with a maximum difference of 1 between declines and improvements.

Regarding the second aim of exploring the predictive value of clinical variables on the postoperative cognitive decline, two levels of analysis were performed. Firstly, participants were classified according to whether they presented a “decline-mixed” or “stable” pattern of cognitive changes. Random forest (RF) regression models were used to assess the differential contributions of the variables that had a higher risk toward the “decline-mixed” pattern of cognitive changes [38]. Secondly, discriminant function analysis was run with the variables that had a higher contribution to present “decline-mixed” pattern of cognitive changes in RF analysis to examine which were the best variables for discriminating between the classification groups. In addition, chi-squared analysis was used to explore association between clinical variables and cognitive pattern.

In cases of missing data, participants were only excluded from the specific analysis for which data were unavailable. They were retained in all other analyses for which complete data were present.

Statistical significance was set at *p*<.05. All analyses were conducted using Jamovi (version 2.6.44). RF analysis and the discriminant function analysis were conducted exclusively in R statistical software, and SPSS-PC software (version 25.0 for Windows), respectively.

## Results

Despite sample loss, the results showed that the baseline CVD group and the HC remained matched in age, sex, years of education, IQ, and handedness preference. In the comparison between the CVD group at follow-up and HC, significant differences were found only in age (*p*=.026), as expected, given the one-year interval. Pre-post comparisons of CVD also revealed significant differences in age (*p*<.001), as well as in the MMSE score (*p*=.034), with lower performance observed one year after surgery. Table 1 shows the sociodemographic, clinical, and general cognitive characteristics of the follow-up study sample.

### Aim 1 – Postoperative changes

Patients with CVD were classified according to the diagnostic criteria of MCI. At baseline, 12/22 patients (54.5%) were classified as CVD-nMCI, while 10/22 (45.5%) presented CVD-MCI. At follow-up, 11/12 patients classified as CVD-nMCI at baseline (91.7%) remained stable in this diagnostic category, while 1/12 participants (8.3%) progressed to CVD-MCI. All patients classified as CVD-MCI at baseline (100%) maintained their diagnosis at follow-up, and none of them developed dementia (Figure 2). Overall, these results demonstrate high diagnostic stability in the sample, with a notably low conversion rate to CVD-MCI (4.5%).

**Figure 2.**
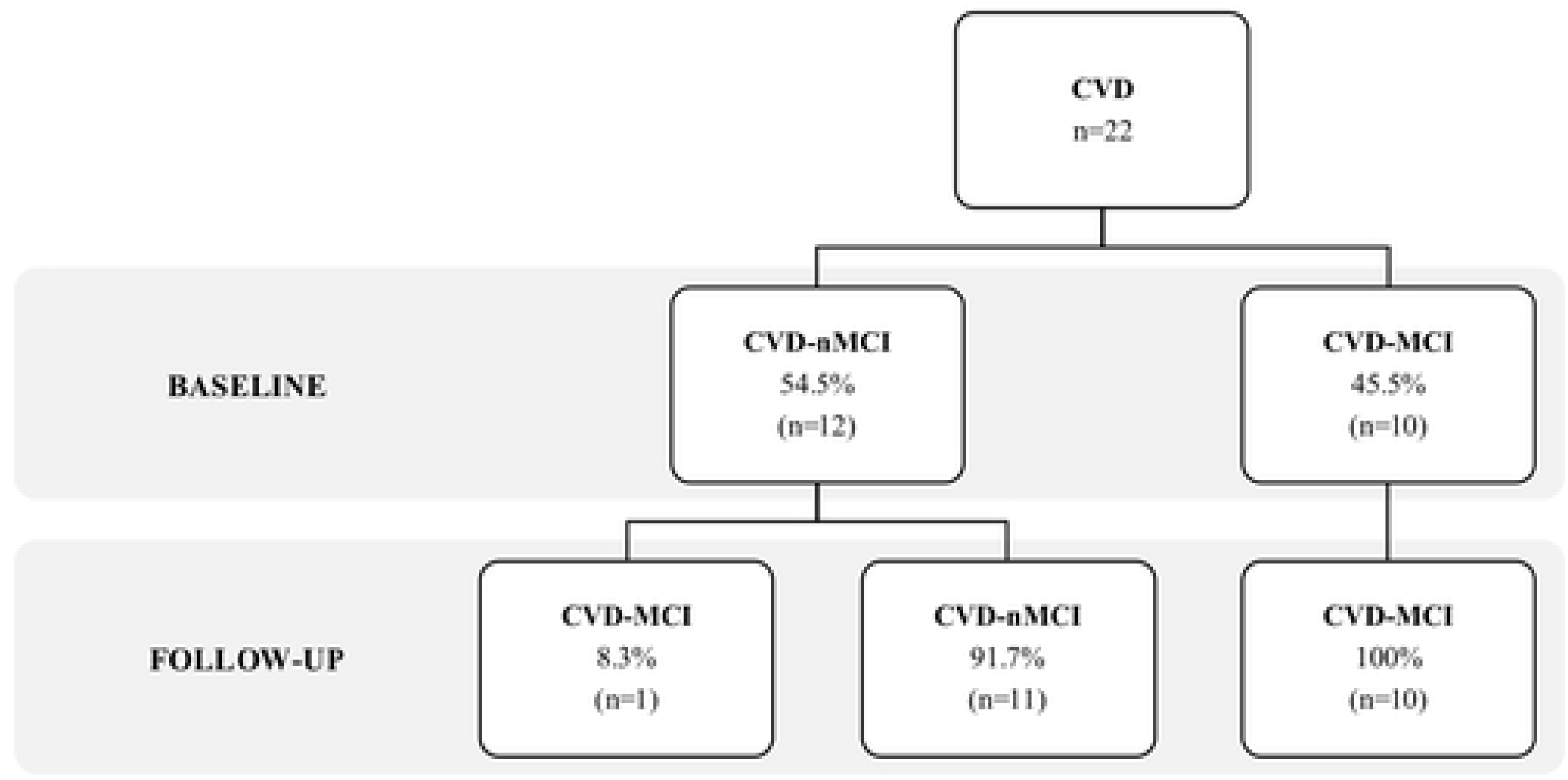
Diagnostic evolution of MCI in participants with CVD between baseline and postsurgical follow-up. Note. Abbreviations: CVD= participants with cardiac disease; CVD-nMCI= CVD participants without Mild Cognitive Impairment; CVD-MCI= CVD participants with Mild Cognitive Impairment; MCI= Mild Cognitive Impairment; n= number of individuals of each group.

The Wilcoxon signed-rank test was used to analyze postoperative changes in cognitive performance in the preoperative MCI subgroups (CVD-MCI and CVD-nMCI). Neuropsychological baseline and follow-up data are presented in Table 3. CVD-MCI showed significant declines in backward digit span, semantic fluency, delayed recall of visual reproduction, and the Verbs Naming Test. CVD-nMCI, showed significant declines in learning and delayed recall of CVLT. No other significant differences were found across cognitive measures.

**Table 3.**
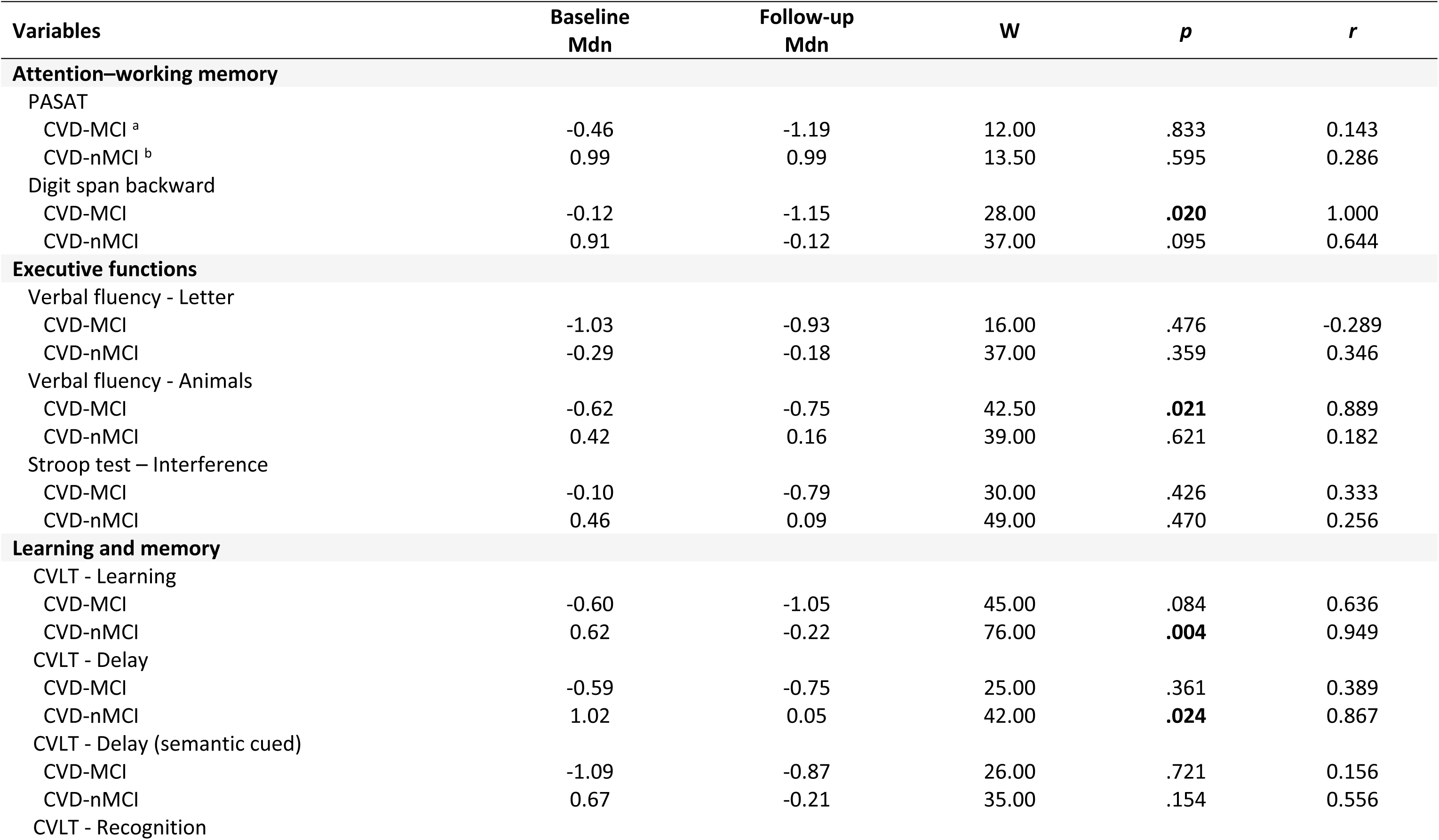
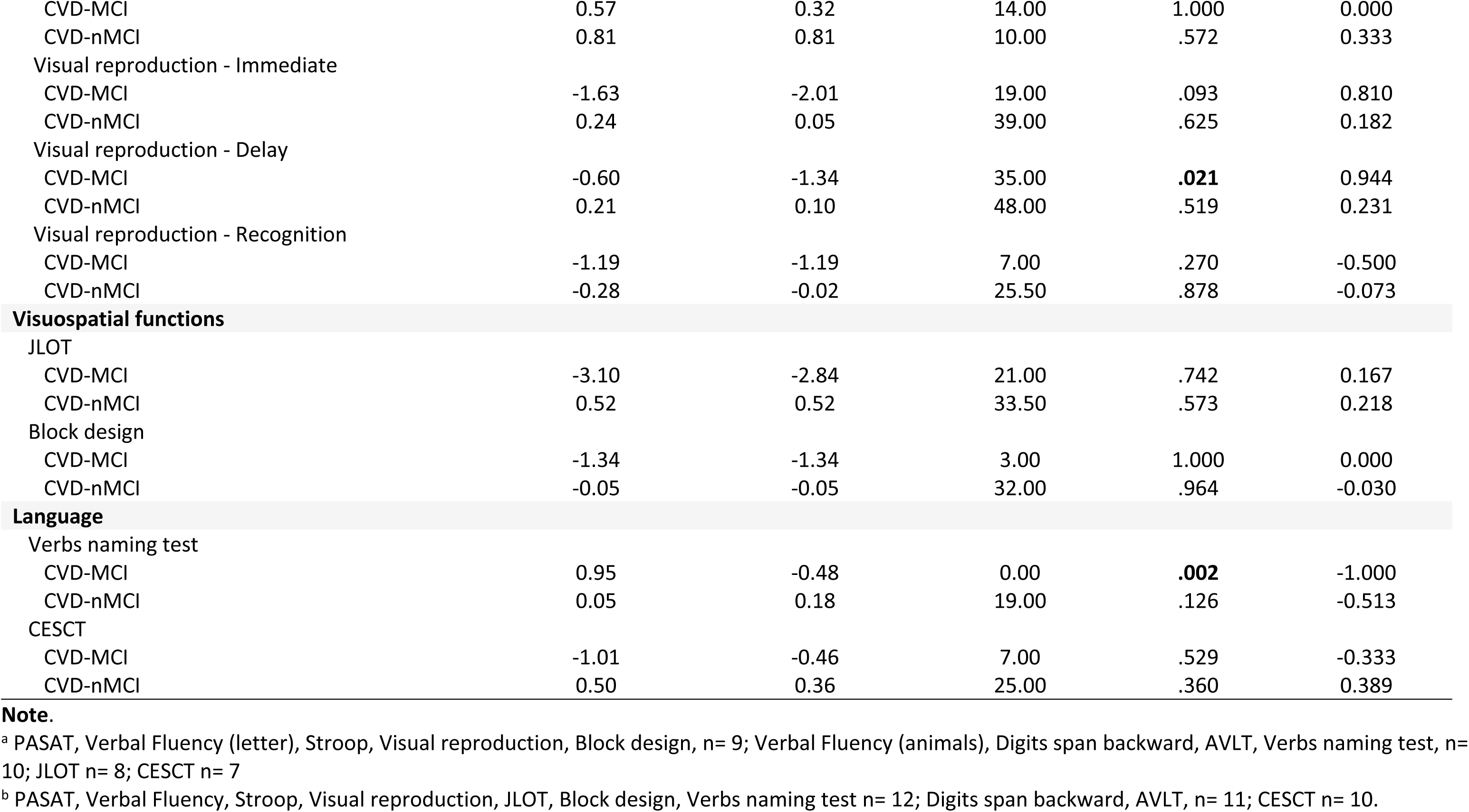
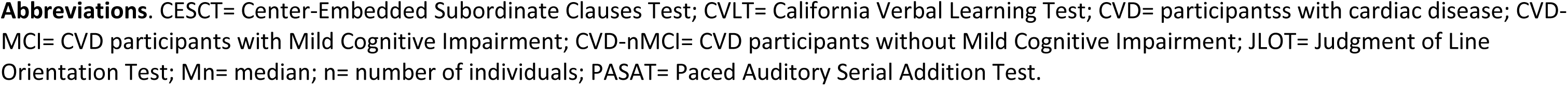
Changes in post-surgical cognitive performance in CVD and pre-surgical MCI diagnostic subgroups.

| Variables | Baseline<br>Mdn | Follow-up<br>Mdn | W | p | r |
| --- | --- | --- | --- | --- | --- |
| **Attention–working memory** |  |  |  |  |  |
| PASAT |  |  |  |  |  |
| CVD-MCI <sup>a</sup> | -0.46 | -1.19 | 12.00 | .833 | 0.143 |
| CVD-nMCI <sup>b</sup> | 0.99 | 0.99 | 13.50 | .595 | 0.286 |
| Digit span backward |  |  |  |  |  |
| CVD-MCI | -0.12 | -1.15 | 28.00 | **.020** | 1.000 |
| CVD-nMCI | 0.91 | -0.12 | 37.00 | .095 | 0.644 |
| **Executive functions** |  |  |  |  |  |
| Verbal fluency - Letter |  |  |  |  |  |
| CVD-MCI | -1.03 | -0.93 | 16.00 | .476 | -0.289 |
| CVD-nMCI | -0.29 | -0.18 | 37.00 | .359 | 0.346 |
| Verbal fluency - Animals |  |  |  |  |  |
| CVD-MCI | -0.62 | -0.75 | 42.50 | **.021** | 0.889 |
| CVD-nMCI | 0.42 | 0.16 | 39.00 | .621 | 0.182 |
| Stroop test – Interference |  |  |  |  |  |
| CVD-MCI | -0.10 | -0.79 | 30.00 | .426 | 0.333 |
| CVD-nMCI | 0.46 | 0.09 | 49.00 | .470 | 0.256 |
| **Learning and memory** |  |  |  |  |  |
| CVLT - Learning |  |  |  |  |  |
| CVD-MCI | -0.60 | -1.05 | 45.00 | .084 | 0.636 |
| CVD-nMCI | 0.62 | -0.22 | 76.00 | **.004** | 0.949 |
| CVLT - Delay |  |  |  |  |  |
| CVD-MCI | -0.59 | -0.75 | 25.00 | .361 | 0.389 |
| CVD-nMCI | 1.02 | 0.05 | 42.00 | **.024** | 0.867 |
| CVLT - Delay (semantic cued) |  |  |  |  |  |
| CVD-MCI | -1.09 | -0.87 | 26.00 | .721 | 0.156 |
| CVD-nMCI | 0.67 | -0.21 | 35.00 | .154 | 0.556 |
| CVLT - Recognition |  |  |  |  |  |
| CVD-MCI | 0.57 | 0.32 | 14.00 | 1.000 | 0.000 |
| CVD-nMCI | 0.81 | 0.81 | 10.00 | .572 | 0.333 |
| Visual reproduction - Immediate |  |  |  |  |  |
| CVD-MCI | -1.63 | -2.01 | 19.00 | .093 | 0.810 |
| CVD-nMCI | 0.24 | 0.05 | 39.00 | .625 | 0.182 |
| Visual reproduction - Delay |  |  |  |  |  |
| CVD-MCI | -0.60 | -1.34 | 35.00 | **.021** | 0.944 |
| CVD-nMCI | 0.21 | 0.10 | 48.00 | .519 | 0.231 |
| Visual reproduction - Recognition |  |  |  |  |  |
| CVD-MCI | -1.19 | -1.19 | 7.00 | .270 | -0.500 |
| CVD-nMCI | -0.28 | -0.02 | 25.50 | .878 | -0.073 |
| **Visuospatial functions** |  |  |  |  |  |
| JLOT |  |  |  |  |  |
| CVD-MCI | -3.10 | -2.84 | 21.00 | .742 | 0.167 |
| CVD-nMCI | 0.52 | 0.52 | 33.50 | .573 | 0.218 |
| Block design |  |  |  |  |  |
| CVD-MCI | -1.34 | -1.34 | 3.00 | 1.000 | 0.000 |
| CVD-nMCI | -0.05 | -0.05 | 32.00 | .964 | -0.030 |
| **Language** |  |  |  |  |  |
| Verbs naming test |  |  |  |  |  |
| CVD-MCI | 0.95 | -0.48 | 0.00 | **.002** | -1.000 |
| CVD-nMCI | 0.05 | 0.18 | 19.00 | .126 | -0.513 |
| CESCT |  |  |  |  |  |
| CVD-MCI | -1.01 | -0.46 | 7.00 | .529 | -0.333 |
| CVD-nMCI | 0.50 | 0.36 | 25.00 | .360 | 0.389 |

**Note.**
<sup>a</sup> PASAT, Verbal Fluency (letter), Stroop, Visual reproduction, Block design, n= 9; Verbal Fluency (animals), Digits span backward, AVLT, Verbs naming test, n= 10; JLOT n= 8; CESCT n= 7
<sup>b</sup> PASAT, Verbal Fluency, Stroop, Visual reproduction, JLOT, Block design, Verbs naming test n= 12; Digits span backward, AVLT, n= 11; CESCT n= 10.
**Abbreviations.** CESCT= Center-Embedded Subordinate Clauses Test; CVLT= California Verbal Learning Test; CVD= participantss with cardiac disease; CVD-MCI= CVD participants with Mild Cognitive Impairment; CVD-nMCI= CVD participants without Mild Cognitive Impairment; JLOT= Judgment of Line Orientation Test; Mn= median; n= number of individuals; PASAT= Paced Auditory Serial Addition Test.

Across cognitive measures, postoperative changes in categorical cognitive status, defined as “decline”, “improvement”, or “stable”, were analyzed in subgroups of preoperative MCI diagnosis (Figure 3, a). CVD-MCI showed notable rates of cognitive decline in attention and working memory, particularly in backward digit span (40%) and PASAT (33.3%). Executive function measures revealed a moderate decline in semantic fluency (20%) and the Stroop interference (22.2%), with some improvements in phonemic fluency (22.2%). In the memory domain, decline was most pronounced in visual reproduction, including immediate recall (22%) and delayed recall (55.6%), followed by verbal memory, including learning (20%) and delayed recall (30%) of CVLT. Some improvements were also observed in memory domain, specifically in CVLT delayed recall with semantic cued (30%), and recognition of visual reproduction (33%). Visuospatial and language functions remained largely stable, with minor improvements, including a 20% improvement in the Verbs Naming Test. McNemar’s test confirmed significant postoperative changes in backward digit span (χ² = 4.000, *p* = .046) and visual reproduction delayed recall (χ² = 5.000, *p* = .025). In contrast, CVD-nMCI showed greater overall stability across cognitive measures (Figure 3, b). In attention and working memory, most participants remained stable, with some decline in backward digit span (27.3%) and to a lesser extent in PASAT (8.1%). Memory and executive functions were largely preserved, with a modest decline in phonemic and semantic fluency (16.7% each), and some improvement in Stroop interference (16.7%). Visuospatial functions were also stable, with some improvement observed in JLOT (16.7%). Finally, language showed moderate improvements, particularly in the Verbs Naming Test (33.3%). No significant postoperative changes were detected in this subgroup using McNemar’s test.

**Figure 3.**
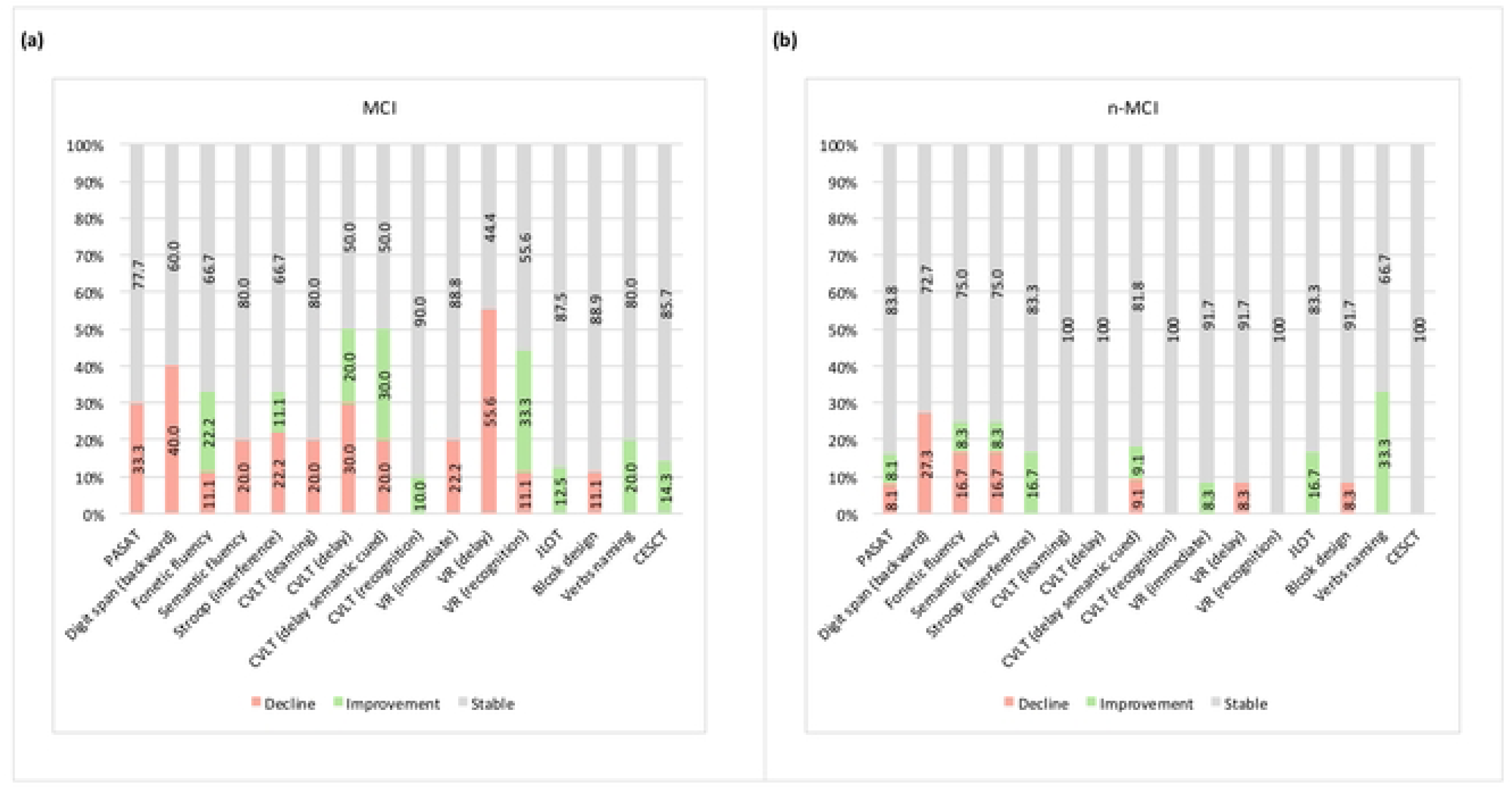
Postoperative changes in categorical status across of cognitive measures in subgroups of preoperative MCI diagnosis.

Intrasubject postoperative cognitive change patterns were analyzed in the CVD-MCI and CVD-nMCI subgroups, based on the number of cognitive variables showing decline, improvement, or stability (Figure 4). In CVD-MCI, 4/10 participants (40%) showed a “decline” pattern, 3/10 (30%) showed a “mixed” pattern, and another 3/10 (30%) remained largely “stable”. In contrast, in CVD-nMCI, 10/12 participants (83.3%) showed stability, 1/12 (8.3%) presented a “decline” pattern, and 1/12 (8.3%) presented a “mixed” pattern. No participants either subgroup showed a predominantly improving pattern.

**Figure 4.**
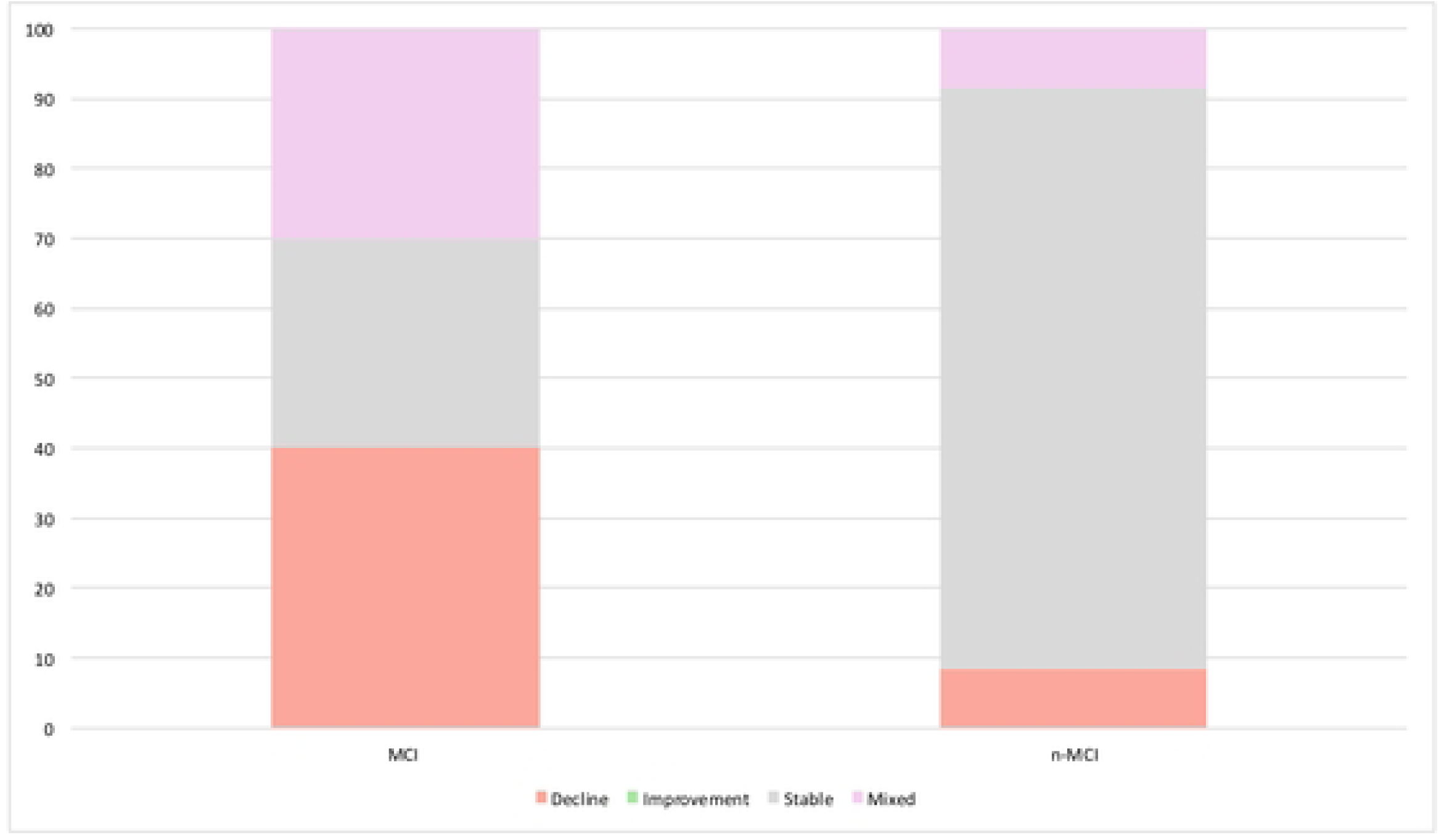
Distribution of postsurgical cognitive change patterns according to presurgical MCI diagnostic subgroups. Note. Abbreviations: CVD-nMCI= CVD participants without Mild Cognitive Impairment at baseline; CVD-MCI= CVD patients with Mild Cognitive Impairment at baseline.

### Aim 2 – Predictors of cognitive changes

The results for aim 1 showed high diagnostic stability of MCI between the preoperative baseline and the postoperative follow-up assessment. Despite this apparent diagnostic stability, the findings also revealed a subclinical and heterogeneous pattern of cognitive decline among participants with CVD-MCI. Considering this, CVD participants were reclassified according to their subclinical pattern of cognitive changes. As a result, 9/22 participants (40.9%) were classified as a “decline-mixed” pattern, whereas 13/22 participants (59.1%) were classified as a “stable” pattern. Preoperative MCI was present in 77.78% of patients in the “decline-mixed” subgroup, compared with 23.07% in the “stable” subgroup (X^2^= 6.42, *p*= .011). The sociodemographic and clinical characteristics of the postoperative cognitive change subgroups are presented in Table 4.

**Table 4.**
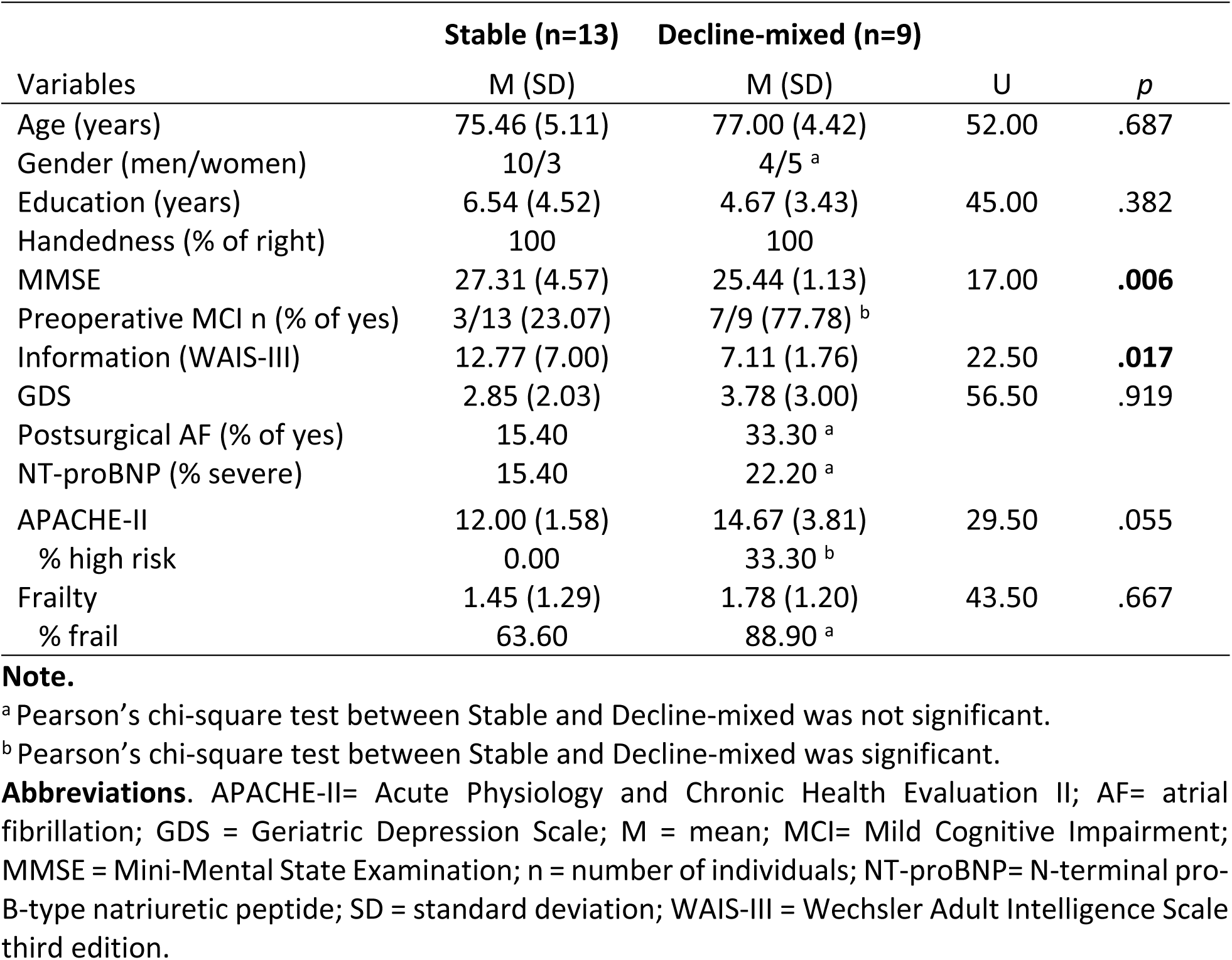
Sociodemographic and clinical characteristics of the postoperative cognitive change subgroups.

For the analysis of aim 2, clinical variables that, according to previous evidence, may be associated with an increased risk of postoperative cognitive decline were selected. The clinical variables entered in the analysis were postoperative atrial fibrillation, NT-proBNP levels, APACHE-II score, and frailty level at 12 months. In addition, sex and estimated IQ were also included. The RF analysis was used to assess the differential contributions of these postoperative variables associated with a higher risk of showed a subclinical pattern of cognitive decline. The RF models showed that IQ, APACHE-II and postoperative frailty were the most important predictors of a “decline-mixed” pattern (Figure 5). Moreover, presence of postoperative atrial fibrillation received a low importance score, while the rest of variables did not contribute to the classification model.

**Figure 5.**
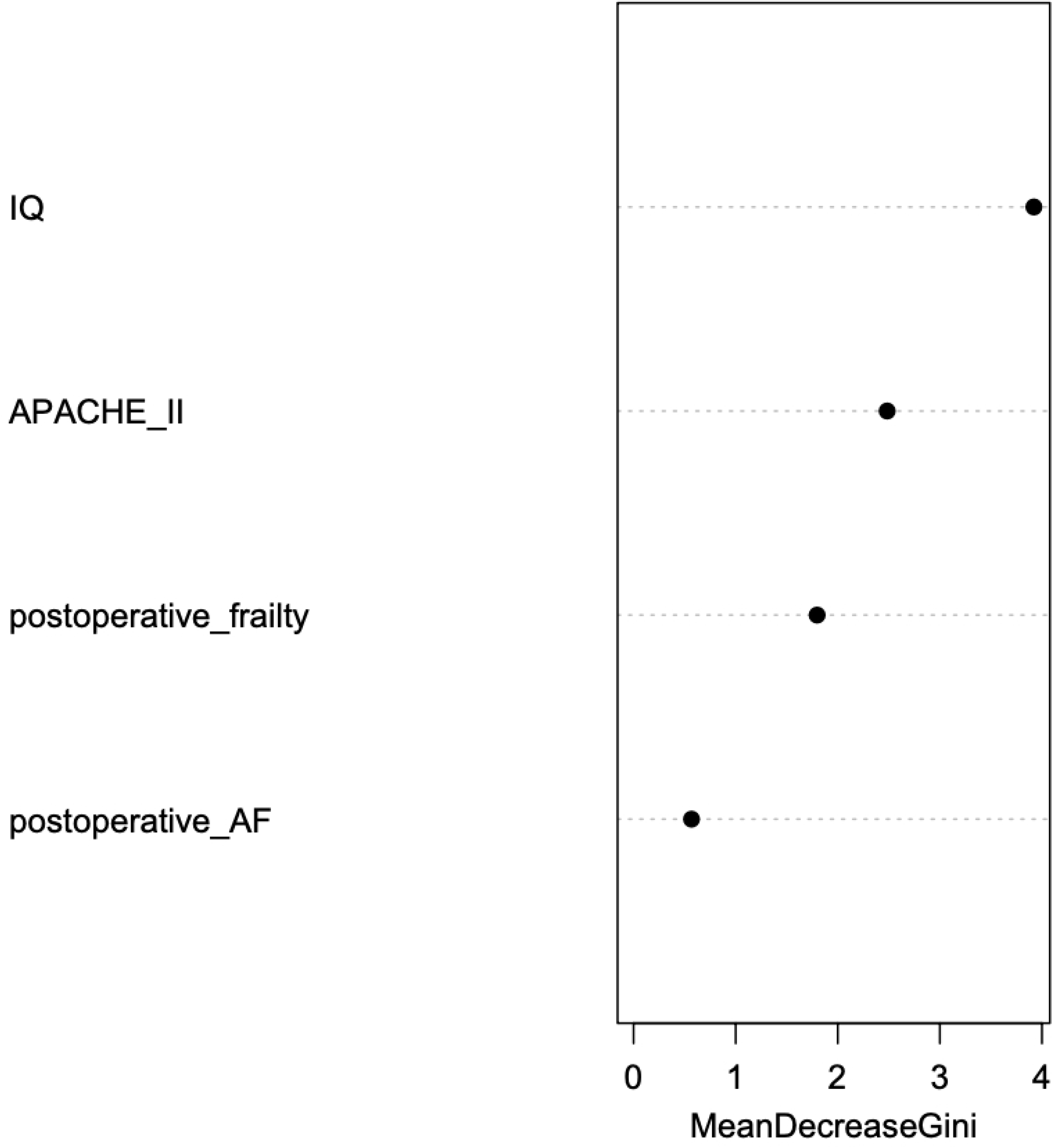
Contribution of postoperative clinical variables to cognitive change patterns. The plot represents the mean decrease in the Gini coefficient; a measure of how each variable contributes to the homogeneity of the nodes and leaves in the resulting RF analysis. A higher mean decrease in the Gini score reflects greater variable importance in the model. Note. Abbreviations: AF = Atrial fibrillation; APACHE-II = Acute Physiology and Chronic Health Evaluation II; IQ= intelligence quotient.

Discriminant function analysis was run to examine which variables best discriminated between groups of cognitive change patterns. Classification of cognitive patterns (decline-mixed, and stable) was included as a dependent variable, while IQ, APACHE-II and postoperative frailty were included as independent variables. The results did not show a significant canonical function (Wilks= .668, X²= 6.664, *p*= .083). However, according to the structure coefficients, IQ (.816) y APACHE-II (-.638) were the strongest discriminators of groups, while frailty (.291) had a lower weight. Regarding the classification of groups, an overall classification rate of 70% was found, with 77.8% of cases correctly classified in the group of “decline-mixed” pattern, and 63.6% of cases in the group of “stable” pattern.

The chi-squared test was used to investigate the association of APACHE-II and postoperative frailty in the patterns of cognitive changes. According to the results, a significant association was found between APACHE-II and “decline-mixed” cognitive pattern (X^2^(1) = 5.02, *p* = .025), while frailty was not significant (X^2^(1) = 1.68, *p* = .194).

## Discussion

The first aim of the present study was to investigate the change pattern in MCI diagnosis and the postoperative neuropsychological profile 12 months after surgery in CVD participants (≥ 65 years). To this end, the rate of change in MCI diagnosis was analyzed, and the clinical characteristics and cognitive performance of the diagnostic subgroups were examined. The results showed that 50% of the participants with CVD met the criteria for postoperative MCI, indicating high diagnostic stability compared with the preoperative frequency of MCI (45.5%). Accordingly, 91.7% of the participants with CVD-nMCI and 100% of those with CVD-MCI at baseline retained their diagnosis at follow-up. Consequently, the conversion rate to MCI was extremely low (1/22, 4.5%) and none of them developed dementia.

To our knowledge, no studies have examined postoperative cognitive status in CVD patients (≥ 65 years) at 12 months after surgery while also applying diagnostic criteria for MCI. Some authors have investigated the presence of cognitive impairment using cut-off scores of screening tests. In this regard, Lingehall *et al.* reported a significant increase in the frequency of cognitive impairment in the early postoperative period, rising from 7.9% preoperatively to 56.1% four days after surgery [12]. However, at the five-year follow-up, these authors observed a decrease in the frequency of cognitive impairment to 26.3%. In contrast, other studies with medium-term follow-up assessments (six weeks and six months after surgery) found no significant differences between preoperative and postoperative cognitive performance in patients with CVD [13,14]. Studies including brief assessment have analyzed cognitive impairment using psychometric approaches. Along this line, Gugino *et al.* defining cognitive impairment as performance ≥ 2 standard deviations below normative data on at least two cognitive measures, found a high frequency of preoperative cognitive impairment (39.5%), with a similar proportion seven days after surgery (40.6%) [15]. Likewise, these authors reported a decrease in the frequency of cognitive impairment to 28% at the three-month follow-up. Newman *et al.* observed a similar pattern, defining cognitive impairment as performance ≥ 1 standard deviation below baseline performance [18]. These authors reported a high proportion of cognitive impairment seven days after surgery (53%), followed by a subsequent decrease at six weeks (36%) and six months (24%). Nevertheless, despite this improvement, they also identified late cognitive decline five years after surgery in 42% of CVD patients. Overall, the available evidence suggests a postoperative course characterized by fluctuations in cognitive status. In the first days after surgery, the proportion of patients with cognitive impairment increases, likely as a transient phenomenon related to the postoperative state and subsequently declines at medium-term follow-up assessments conducted months after surgery. However, longitudinal studies with long-term follow-up (≥ 1 year) have reported the emergence of late cognitive decline in these patients. In this regard, despite the diagnostic stability of MCI observed in the present study, the results reflect a heterogeneous pattern of postoperative changes in CVD patients one year after surgery.

In the present study, the analysis of the MCI diagnostic subgroups showed that, patients with preoperative CVD-MCI exhibited decline in working memory and executive functions, as well as in visual memory and the naming component of language, whereas participants with preoperative CVD-nMCI only showed a significant decline in verbal memory. These findings may reflect an evolution of the preoperative profile described by Mata *et al.* in which patients with CVD-MCI already showed greater impairment in verbal memory and visuospatial functions, which could explain the absence of significant postoperative decline in these functions [10]. By contrast, patients with CVD-MCI showed postoperative worsening in functions that had shown a lesser degree of impairment at the preoperative stage, such as visual memory, executive functions, and the naming component of language. Conversely, patients with CVD-nMCI showed a relatively preserved preoperative profile compared with the CVD-MCI. Therefore, despite postoperative diagnostic stability, the findings of the present study indicate the presence of subclinical cognitive decline in participants with CVD, which appears to be more pronounced in those who had MCI before surgery.

To further characterize this subclinical decline in CVD, the present study analyzed postoperative changes in categorical cognitive status in the cognitive measures of the preoperative MCI subgroups. The results showed predominantly stable performance across all cognitive measures in the preoperative CVD-nMCI. In contrast, the preoperative CVD-MCI showed a more heterogeneous pattern of change, with a higher percentage of declines compared with the CVD-nMCI, particularly in visual memory, in which 55.6% of participants with CVD-MCI showed postoperative decline compared with 8.3% of those with CVD-nMCI (Figure 3). This pattern is further supported by the results of the within-subject analysis, according to which the CVD-nMCI showed a higher proportion of participants following a “stable” pattern (83.3%) and, to a lesser extent, a “decline” (8.3%) or “mixed” pattern (8.3%). By contrast, the CVD-MCI showed greater heterogeneity, with a high proportion of participants following a “decline” pattern (40%), followed by a “mixed” pattern (30%) and a “stable” pattern (30%). Taken together, these findings also suggest greater postoperative cognitive vulnerability in participants with CVD-MCI. Likewise, the postoperative stability of the MCI diagnosis, in contrast to the subclinical changes observed, highlights the need to characterize the neuropsychological profile of patients with CVD, with particular emphasis on the preoperative neuropsychological profile as a prognostic factor in the postoperative evolution of cognitive status.

The second aim of the present study was to explore whether sociodemographic and clinical variables predict postoperative cognitive decline. The results of the RF analysis indicate that estimated IQ, APACHE-II classification score, and postoperative frailty are the most important variables in predicting the postoperative “decline-mixed” pattern. The findings regarding estimated IQ are consistent with those reported in previous studies, in which lower IQ is associated with a greater risk of cognitive impairment in the preoperative stage [10,39]. Given that preoperative cognitive status is related to postoperative outcome, it is reasonable to expect that estimated IQ also contributes significantly to the prediction of postoperative cognitive status. In addition, other authors have investigated different proxies of cognitive reserve, such as educational level, and found that lower levels of this indicator are associated with greater postoperative cognitive impairment five years after surgery [18]. Likewise, in a recent study, Megari and Kosmidis reported a significantly higher incidence of postoperative cognitive decline in CVD patients and low cognitive reserve, whereas those with high cognitive reserve maintained a stable cognitive performance [40]. These findings are consistent with those of other studies describing cognitive reserve as a protective factor against the development of neurodegenerative diseases [41]. Taken together, these results suggest that indicators of cognitive reserve, particularly high IQ, may act as protective factors for cognitive performance in patients with CVD from the preoperative stage onward and as modulators of postoperative cognitive trajectory.

With regard to perioperative variables, the findings of the present study identify the APACHE-II classification score as the second most important variable in the prediction model. This index is considered a marker of illness severity and is commonly used at intensive care unit admission to estimate mortality risk. To our knowledge, no studies have examined the relationship between APACHE-II and postoperative cognitive impairment in patients with CVD, which adds novelty and value to the findings reported here. Despite the absence of studies directly linking these two variables, the available literature has identified a high APACHE score as a risk factor for the development of postoperative delirium [42]. Evidence suggests that postoperative delirium constitutes a risk factor for the development of long-term cognitive impairment [43]. In this context, because APACHE-II reflects greater severity of the patient’s overall clinical condition, it may be considered a marker of cerebral vulnerability that increases the risk of postoperative delirium and, according to the findings of the present study, of long-term cognitive decline.

Frailty status is an important variable in the prediction model, although with less weight than IQ and APACHE-II. In addition, the complementary chi-square analysis showed that postoperative frailty was not significantly associated with the postoperative decline pattern, whereas APACHE-II score did show a significant association. Nevertheless, these findings should be interpreted with caution given the small sample size. These results are also novel, as no published studies were identified that have examined the predictive value of postoperative frailty status in relation to postoperative cognitive impairment in patients with CVD. Again, as with APACHE-II, the available studies investigating frailty in the context of CVD have generally included it as a predictor of postoperative delirium [20]. However, despite the lack of postoperative studies, some authors have examined the association of preoperative frailty with preoperative MCI diagnosis in CVD and found no significant results [37]. In the present study, frailty was assessed using the Fried phenotype, which captures physical frailty. Nevertheless, the concept of frailty has recently evolved toward a more multidimensional approach that includes cognitive and social domains in addition to the physical domain [44]. In this context, instruments such as the Essential Frailty Toolset have been proposed in recent years [45]. This tool may have provided a broader characterization of clinical vulnerability, including cognitive and systemic components, which are particularly relevant in older cardiac patients.

A limitation of the present study is the small sample size; therefore, the findings should be interpreted with caution. Another limitation is the absence of follow-up data for the HC group. Accordingly, follow-up studies with larger samples and an HC group are needed to confirm these findings. Additionally, further research is required to better understand the cognitive heterogeneity of CVD, incorporating comprehensive neuropsychological assessments and neuroimaging data. More studies are also needed to examine the predictive role of relevant clinical variables, such as NT-proBNP levels and postoperative atrial fibrillation, in larger samples. Furthermore, longer-term longitudinal follow-up (> 1 year after surgery) is needed to determine whether cognitive changes observed one year after surgery are associated with a later risk of developed dementia. Nevertheless, despite these limitations, the present study has important clinical value. The findings contribute to a better understanding of cognitive impairment associated with CVD, which may help promote diagnostic strategies aimed at the early detection of Va-MCI and, considering the trajectories of postoperative cognitive change, its progression to vascular or mixed dementia.

## Conclusions

In summary, the present study explores the postoperative cognitive trajectory of older patients with CVD by combining an MCI diagnostic approach with a clinical analysis of postoperative cognitive change patterns, based on a comprehensive neuropsychological assessment conducted before and after surgery. The findings provide a deeper understanding of the nature of cognitive impairment in CVD, indicating a high frequency of MCI both before and after surgery, with notably similar rates across the two time points. Despite this diagnostic stability, the results indicate the presence of subclinical cognitive decline in patients with CVD, following a heterogeneous pattern that appears to be more pronounced in those who had MCI before surgery. In addition, important variables were identified in the prediction of postoperative cognitive impairment, including IQ, APACHE-II score, and postoperative frailty, which may contribute to the identification of particularly vulnerable patients and help guide preventive interventions tailored to each patient’s profile. These findings are of considerable value for future research and clinical practice, given the increase in life expectancy and the high incidence of CVD in older adults.

## Data Availability

The minimal data set is available at Zenodo via https://doi.org/10.5281/zenodo.20643848

## Acknowledgments

The authors would like to express their gratitude to Dr. Rafael Martínez Sanz, Head of the Cardiac Surgery Department of the University Hospital of Canary Islands, and all the cardiac surgeons of this department, as well as Dr. Elena Sirumal Rodríguez for their support in the recruitment. The authors would also like to extend their appreciation to Patrick Dennis for his English language editing support.

